# The spread of a validated molecular marker of artemisinin partial resistance *pfkelch13* R622I and association with *pfhrp2/3* deletions in Eritrea

**DOI:** 10.1101/2023.10.20.23297302

**Authors:** Selam Mihreteab, Karen Anderson, Irene Molina - de la Fuente, Colin J. Sutherland, David Smith, Jane Cunningham, Khalid B Beshir, Qin Cheng

## Abstract

Eritrea is the first African country to switch away from exclusive use of HRP2-based RDTs for the detection of *P. falciparum* due to high prevalence of *pfhrp2*/*3-*deleted *P. falciparum* parasites causing false-negative RDT results. While heavy reliance on malaria RDTs played a significant role in the rapid expansion of *pfhrp2/3*-deleted parasites in Eritrea, we hypothesize that the use of antimalarial (artesunate-amodiaquine) may have also contributed to their spread. We conducted a retrospective investigation of mutations in the propeller domain of the *P. falciparum kelch13* gene in samples collected in 2016 (n=50) from the Northern Red Sea Zone before the RDT switch away from HRP2-RDTs and in samples collected in 2018-2020 (n=587) from the Gash Barka, Anseba and Debub Zones after the RDT switch. No mutations were identified in the 2016 samples. However, in 2018-2019 samples, we detected five different single non-synonymous mutations. The most prevalent mutation was *pfk13* R622I, which was detected in samples collected from all health centres, with an overall prevalence of 11.9% (ranging from 5.9% to 28%). Parasites carrying the R622I mutation have diverse microsatellite marker haplotypes, indicating that they had evolved multiple times from different genetic backgrounds. The prevalence *pfk13* R622I was significantly higher in single *pfhrp3*-deleted parasites (18.0%) compared to parasites without *pfhrp2/3* deletions (6.2%) and dual *pfhrp2/3*-deleted parasites (9.0%), suggesting association between the *pfk13* R622I mutation and the *pfhrp2/3* deletions in Eritrea. Continuous monitoring the trends in *pfhrp2/3* and *pfk13* mutants is needed to inform effective malaria management strategies in Eritrea.

## BACKGROUND

Eritrea, located in the Horn of Africa, experiences low and seasonal transmission of both *P. falciparum* and *P. vivax* malaria^1^. In 2012, the Eritrean National Malaria Control Programme (NMCP) successfully implemented a range of malaria interventions, including integrated vector control, the introduction of quality malaria RDT, and the deployment of artemisinin combination therapy (ACT)^2^. However, between 2015-2020, malaria incidence and mortality rates increased by more than 40%^1^. This resurgence can be attributed, in part, to the emergence of mutant parasites capable of evading diagnostic tests and the development of partial resistance to artemisinin drugs among the parasites^3^.

In 2006, Eritrea introduced a HRP2-based combo RDT as a diagnostic tool for case malaria management. However, within the span of a decade, the country reported a high prevalence (62.0%) of *P. falciparum* parasites carrying *pfhrp2/3* gene deletions causing high rates of false negative HRP2-RDT results among symptomatic patients^4,5^, making Eritrea one of the countries severely impacted by these *pfhrp2/3*-deleted parasites. As a result, in 2016, Eritrea switched from HRP2-based RDTs to non-HRP2-based alternatives^6^. In 2019, approximately 2.5 years after the RDT switch, while the prevalence of parasites with gene deletions remains high the prevalence of parasites with dual *pfhrp2/3* deletions, a primary contributor to false negative RDT results, was much lower at multiple survey sites compared to that at the original sites^6^.

In 2007, following the introduction of RDTs, the Eritrean NMCP made a decision to switch the first line treatment of uncomplicated *P. falciparum* malaria from chloroquine and sulphadoxine/pyrimethamine (CQ-SP) to the ACT artesunate-amodiaquine (AS-AQ) due to a high treatment failure rate associated with CQ-SP^7^. To monitor the efficacy of AS-AQ, as recommended by WHO, therapeutic efficacy studies (TES) have been conducted within the country. These studies, conducted at various sentinel sites, have consistently demonstrated that AS-AQ exhibited efficacy rates exceeding 90% on day 28 in all sites^8–10^, however, the patient Day-3 parasite positive rate has increased between 2016 and 2019^10^.

In addition to TES, surveillance was undertaken to monitor molecular markers associated with partial artemisinin resistance, point mutations in *P. falciparum kelch13* gene (*pfk13*), as well as resistance to CQ (mutations in *pfcrt*) and AQ (mutations in *pfcrt* and *pfmdr1*). These surveillance activities were conducted in the Gash Barka, Medefera and Debub Zones between 2013-2014^11,12^ and in Gash Barka and Northern Red Sea Zones between 2016-2019^10^. The findings from these surveillance efforts supported the TES results. While high prevalence of AQ resistance-associated alleles of *pfcrt* (CVIET haplotype) and *pfmdr1* (Y184F and N86Y) were continuously detected, prevalence of *pfk13* mutations was low in 2013-2014, with a single R622I and T535M detected among 119 samples from Gash Barka, as determined through deep amplicon sequencing. However, a recent report showed that the prevalence of *pfk13* R622I mutation in isolates before treatment reached 8.6% in 2016 and increased to 21.0% in 2019^10^.

The *pfk13* R622I mutation was first reported in a single sample collected from Zambia during the years 2012-2014, as part of a broader study on the global distribution of *pfk13* mutations^13^. Subsequently, it was reported in two individual samples collected from Mozambique and Somalia^14^. In Northern Ethiopia, specifically in a town close to the border with Sudan, the prevalence of R622I was found to be 2.4% (3/25) in samples collected during 2013-2014^15^. This prevalence increased to 9.3% (8/86) in samples collected between 2017 and 2018, indicating an expansion of the mutant in the area^16^. The R622I mutation has been shown to possibly affect the function of *pfk13* by molecular modelling^15^, and it has recently been reported to associate with higher risk of day-3 positivity and recrudescence following ACT treatment in Eritrea TES^10^. In gene-edited parasites, the R622I mutation confers low levels of resistance to artemisinin in vitro^10^.

While existing data suggest that the major driving force behind the high prevalence of *pfhrp2/3* deletions in Eritrea was likely due to the use of HRP2-based RDTs ^6^, it is important to consider that several other factors might also contribute to the rapid increase of the parasites in the country. Theoretically, various mechanisms such as fitness gains, the development of drug resistance, and the absence of host immunity could all confer selective advantages to *pfhrp2/3*-deleted parasites when facing competition or encountering pressures from drug and immune system from the host. Conversely, the evasion of detection by RDTs due to *pfhrp2/3* deletions could also contribute to the survival and spread of drug resistant parasites. Consequently, *pfhrp2/3*-deleted parasites could spread in communities by hitch-hiking alongside the spread of drug-resistant parasites or a newly emerging strains for which there is no pre-existing immunity in the community^17^. Conversely, drug resistant parasites might be similarly spread by hitch-hiking on the spread of *pfhrp2/3*-deleted parasites. The interplay between *pfhrp2/3*-deleted parasites and drug-resistant parasites is not clear, with both phenomena potentially influencing each other’s spread and survival within communities.

In this study, we conducted a retrospective analysis *pfk13* mutations using existing samples collected from symptomatic patients seeking care more than 10 health facilities across 3 Zones as part of previous studies. These samples were originally collected during three separate periods: during 2016 survey and the 2019-2020 survey, both surveys aimed at assessing prevalence of *pfhrp2/3* deletions; and during the 2018-2019 severe malaria study. Our primary objective was to investigate the prevalence of *pfk13* mutations in Eritrea and explore their potential relationship with *pfhrp2/3* deletions. The study contributes valuable insights into the potential selective forces the influence the relative abundance of diagnostic and drug-resistant parasites in the Eritrean context.

## METHODS

### Source and number of samples

In this retrospective study, patient samples were collected from two different studies:

1. Surveillance of *pfhrp2/3* deletions: This study incorporated samples obtained from two cross-sectional surveys designed to investigate the prevalence of *pfhrp2/3* deletion among symptomatic patients (referred to as “survey samples”). The first survey was conducted in 2016 at two hospitals in the Red Sea Zone, namely Ghindae and Massawa^4^, from which 50 samples were obtained. A follow up pfhrp2/3 deletion survey took place during 2019-2020, encompassing nine hospitals/Health Centres spanning three zones: Gash Baka (Agordat, Shambuko, Tesseney, Tekobmia and Barentu), Anseba (Keren and Hagaz) and Debub (Mendefera and Maimine)^6^. This boarder survey collected 714 *P. falciparum* samples, with 417 (representing 66.0%) of these samples were used in the present study.
2. The severe malaria study. The study was conducted during the period of 2018-2019 with the aim of investigating the impact of *pfhrp2/3* deletions on clinical outcomes in Eritrea. A total of 116 *P. falciparum* infected blood samples (referred to as “severe malaria samples”) were collected from three distinct zones: Gash Barka (Agordat and Barentu), Anseba (Keren) and Debub (Menderfera, Emni-Haily, Areza, Dubarwa, and Mulki).

For data analysis, survey and severe malaria samples, collected between 2018 and 2020, were merged according to health facilities where they were collected. In cases where less than five samples were obtained from each of the four health facilities in the Debub zone (Areza, Dubarwa, Emni-Haily and Mulki), these were consolidated into a single group (A/D/E/M). The collection sites can be visualised on the map provided Figure 1.

**Figure 1.**
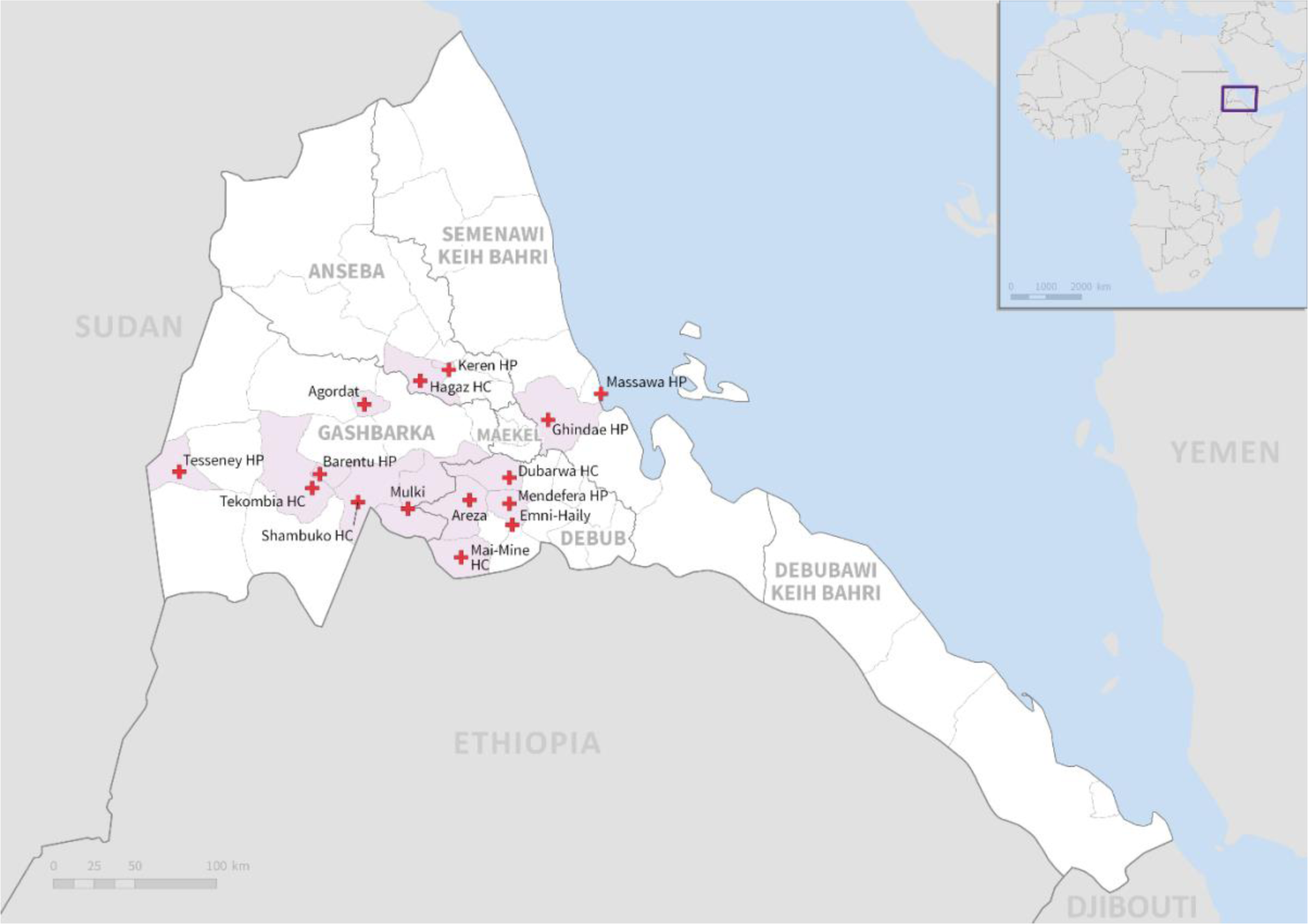
Map of Eritrea showing health facilities where samples were collected.

### Genomic DNA and Plasmodium infection

Genomic DNA extraction from survey samples had been previously described in earlier publications^4,6^. Additionally, genomic DNA were isolated from severe malaria samples using robotic system, following the same protocol as previously reported^18^.

### Amplification and sequencing *pfk13*

For survey samples, which included 50 samples from the 2016, and 417 DNA samples randomly selected from different *pfhrp2/3* status groups of 2019-2020, the propeller region of the *pfk13* gene (codons 435 to 680) was amplified using published primers and conditions^19^. Subsequently, amplified fragments were then sequenced from both ends by Sanger sequencing using BigDye reagents.

For severe malaria samples, A 1033bp fragment, spanning from codon 380 to 724 of the propeller region of *pfk13* amino acid sequences, was amplified using forward primer (TAAGTGGAAGACATCATGTAACCAGAGA) and reverse primer (GTCTAAAACCAGTGGAACAAAAATCGT). Amplification of DNA was performed in a 25 µl reaction on thermal cycler (PCRmax, UK). The reaction mixture included 5 µl of extracted genomic DNA, 200 nM of each primer, 3 mM of MgCl2, 300 μM of each dNTP (Meridian, UK), 1X NH4 reaction buffer (Meridian, UK) and 1 U of the NEB High Fidelity Q5 polymerase. The PCR conditions consisted of 95°C for 3 min, followed by 40 cycles of denaturation at 95°C for 30 sec, annealing at 55°C for 60 sec, and extension at 72°C for 2 min. The amplified fragments were visualised using gel electrophoresis^18^. Subsequently, amplicon pooling and cleaning were carried out as previously described^20^. Library preparation and adaptor ligation were carried out using the Oxford Nanopore native barcoding kit (sqk-nbd-112-96) in accordance with the manufacturer’s protocol (Oxford Nanopore, UK).

### Haplotype diversity and genetic relatedness

A total of seven neutral microsatellite markers (TA1, PolyA, PfPK2, TA109, 2490, 313, and 383) were analysed as part of previous studies^4,6^. Haplotype data were available for 184 survey samples, which were included in the current study - 50 from 2016 and 134 from 2019/2020. To assess the genetic diversity and genetic relatedness of parasite haplotypes with different *pfk13* variants, we employed FSTAT and PHYLOViZ online software (version 1.1) using instructions provided by the developers.

### *pfhrp2* and *pfhrp3* gene status

The status of *pfhrp2* and *pfhrp3* gene in all samples was evaluated using a multiplex qPCR method. This method simultaneously amplifies a fragment of the *pfhrp2* and *pfhrp3* exon2, pLDH and human tubulin gene ^21^. Published primer and probe sequences, as well as PCR reagents and conditions, were used for assessing sever malaria samples. For survey samples, we introduced some modifications: quencher on the *pfhrp2* (BHQ1) and *pfldh* (BHQ2) probes were adjusted, Quantinova multiplex PCR kit master mix (QIAGEN) was utilized, and the Mic qPCR cycler (Bio Molecular Systems) was employed. In the classification of samples as having a pfhrp2 and/or pfhrp3 deletion, a threshold was set at ΔCq (Cq_pfhrp2_ - Cq_pfldh_ and/or Cq_pfhrp3_ - Cq_pldh_) ≥ 3^21^. Samples exhibiting Cq_pldh_ values exceeding 35 cycles were deemed indeterminate due to insufficient DNA for analysis.

### Quantification of parasite density

To determine the quantity of parasite DNA (pg/µL) in survey samples, we relied on the Cq_pldh_ value of each sample in comparison to a 3D7 standard curve. This standard curve was generated using serial dilutions of 3D7 DNA at 1, 0.1, 0.01 and 0.001ng/µL, and it was included in each multiplex qPCR run. In case of the severe malaria samples, parasite density estimation was conducted against the WHO International Standard^22^, as previously described^18^.

### Ethics considerations

The surveys of *pfhrp2/3* deletions were granted approval by the Eritrean MOH Research and Ethical Committee. Laboratory analyses on collected samples received approval from the Departments of Defence and Veterans Affairs Human Research Ethics Committee (DDVA HREC 15-004). Additionally, the severe malaria study was granted approval by LSHTM Ethics Review Committee (#11979).

### Data analysis

Sequence analysis: Sequences of *Pfk13* from survey samples were aligned using MEGA7.0.26^23^. SNPs were identified by comparing these alignments with *pfk13* sequence in 3D7 (PF3D7_1343700). For severe malaria samples, amplicon reads were analysed using the python language and well-established bioinformatics tools as described previously, with some slight modifications^24^. Sequencing alignment was performed using minimap2 software^25^. Mutation prevalence analysis was conducted using the R-package vcfR (version 1.12.0) and plyr (version 1.8.6).

Statistical analysis: To compare mutation prevalence and proportions between sample groups, chi-squired test and odds ratio (OR) analysis were employed. Additionally, comparisons of median parasite DNA concentrations and parasite densities among sample groups were carried out using Mann-Whitney test.

## RESULTS

### Prevalence of *pfk13* mutation

Sequence analysis of 50 samples collected in 2016 revealed a wild type *pfk13* gene in all samples, indicating the absence of mutations. In contrast, analysis of sequences obtained from the combined pool of 587 survey and severe malaria samples collected between 2018 and 2020) samples revealed five different non-synonymous mutations: E605K, R622I, N657K, K658E and S679L. Importantly, each mutant *pfk13* sequence contain only a single amino acid mutation within the propeller region, with no combinations observed. Among these mutations, the R622I mutation had the highest overall prevalence of 11.9% (70/587). Following this, N657K the N657K mutation was observed in 0.3% of the samples (2/471). Meanwhile, the E605K, K658E and S679L mutations were each detected in a single sample, giving a 0.2% (1/587) prevalence (Supplemental Table 1).

### Distribution of *pfk13* mutations

The samples carrying the E605, K658E, S679L and N657K mutations were exclusively collected from the Gash Barka zone. However, the R622I mutation was identified in samples collected from all health facilities in the three zones with a prevalence exceeding higher than 10% in eight of these health facilities (Figure 2, Supplemental Table 1). The prevalence of R622I was highest in Hagaz hospital (28.0%, 7/25) in the Anseba zone, while the lowest prevalence was observed in samples obtained from Mendefera Hospital (5.9%, 3/51) in the Debub zone. The prevalence of R622I was significantly higher in Hagaz compared to Mendefera (Chi-squared test, p=0.0078), Keren (9.9%, Chi-squared test p=0.0207) and Barentu (p=0.0341). However, no statistical differences were observed at the zone level (p>0.05). The prevalence for R622I were 11.9% (44/370) in Gash Barka, 13.8% (16/116) in Anseba and 9.9% (10/101) in Debub zone (Figure 2, Supplemental Table 1).

**Figure 2.**
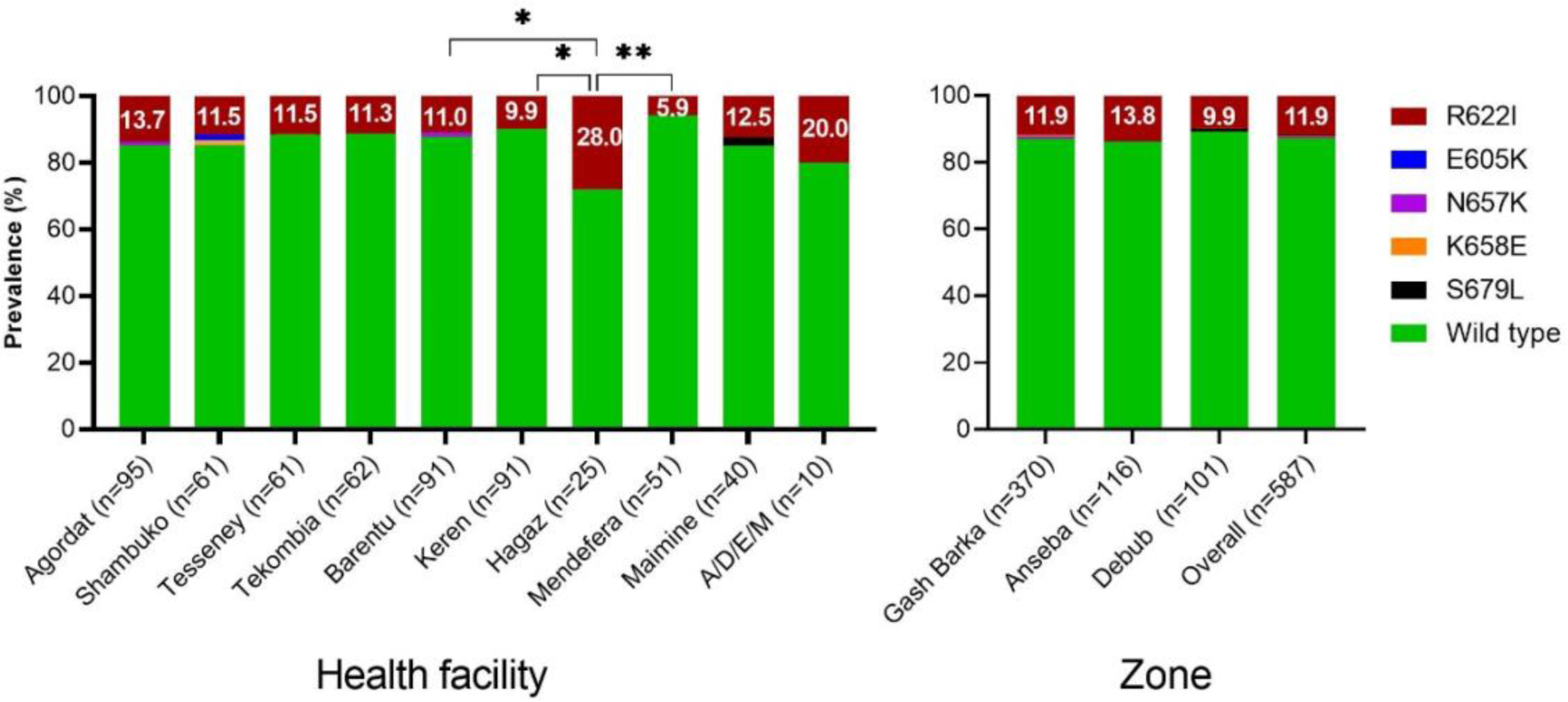
Prevalence of K13 mutations among health facilities, zones and overall. *p<0.05; **p <0.01.

### Haplotype diversity and genetic relatedness of *pfk13* R622I mutants

To gain insight into whether the R622I mutants shared a common ancestry and to assess if they were undergoing expansion, we analysed microsatellite haplotypes obtained from previous studies^4,6^. We examined 184 survey samples, of which 26 carried the *pfk13* R622I mutation. Among these 26 samples, we identified 23 unique haplotypes, with all but two were found in one sample each. The remaining two haplotypes were shared between two and three samples (Supplemental Table 2). In contrast, among the remaining 158 samples with the wild-type 622R, we identified 87 unique alleles. The overall genetic diversity, represented by H*_E_* values, among R622I mutants was calculated at 0.4774 (95% CI: 0.1873 – 0.7676), which is slightly lower than that of the 622R wild-type parasites (0.5773, 95% CI: 0.3760 – 0.7786). However, these difference was not statistically significant (FSTAT, p=0.0500, determined after 20 permutations, Supplemental Table 3).

Our genetic relatedness analysis revealed that while 15 of the 23 R622I mutant haplotypes were closely related within a cluster, the remaining eight mutant haplotypes were not closely related to each other, suggesting independent genetic origins (Figure 3). These findings indicate parasites carrying R622I mutation had evolved from different genetic backgrounds, involving both clonal expansion and independent emergence.

**Figure 3.**
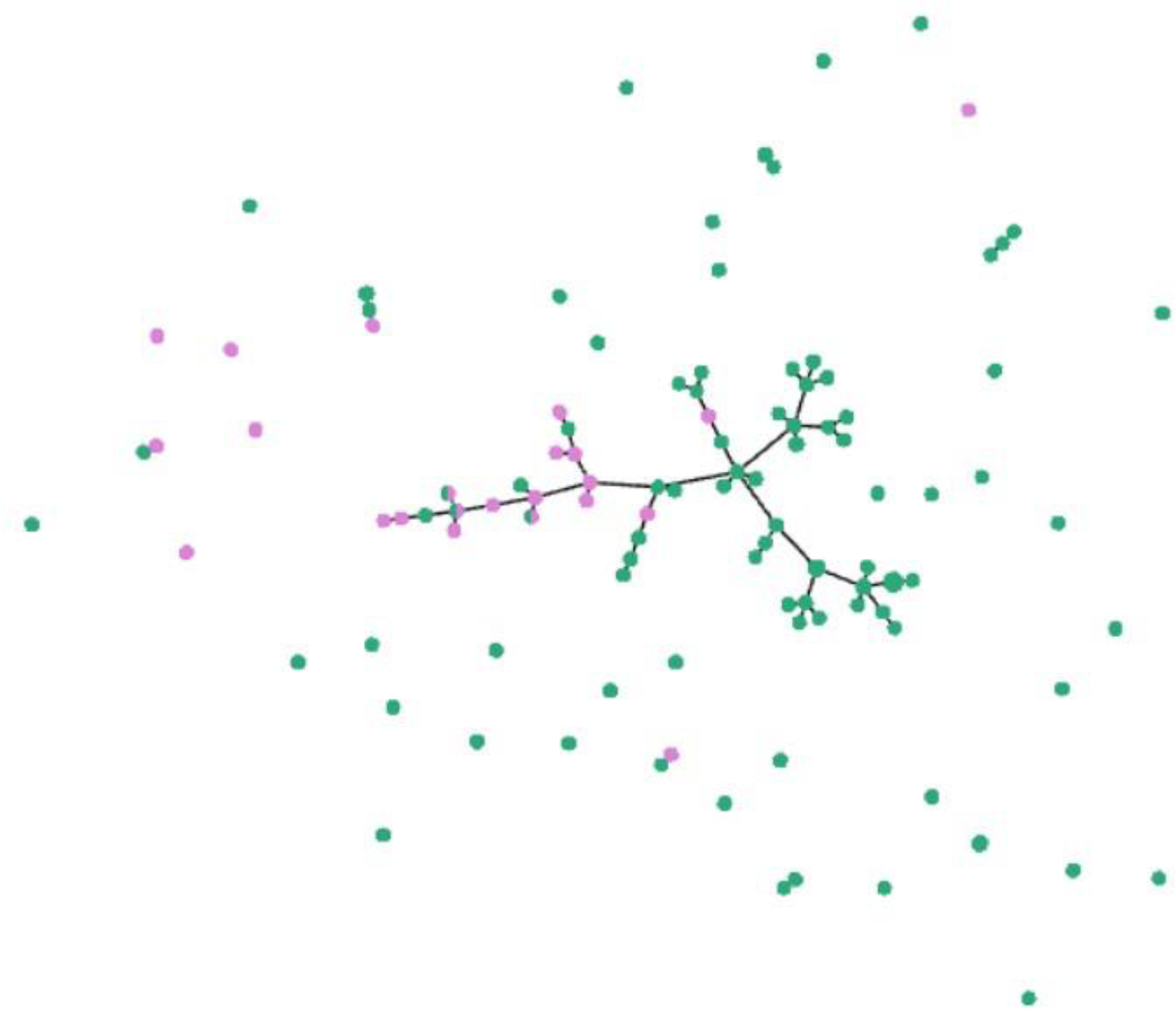
Genetic relatedness of parasites carrying the *pfk13* R622I mutation (in pink) or wild type 622R (in green). Cut-off of 2 was used, in that only parasite haplotypes with ≥ 5 identical alleles of the 7 markers tested were linked together.

### Parasite density and *pfk13* mutations

To investigate whether the *pfk13* mutations had impact on parasite fitness, we analysed the parasite densities estimated for the survey samples, and we found no significant difference in median parasite DNA concentration between samples carrying the R622I mutation and those with the wild-type 622R (25.15, 95%CI: 18.55 – 40.72 vs 23.71, 95%CI: 18.55 – 28.23, p=0.6771, Figure 4). Similarly, our analysis of severe malaria samples indicated comparable (p=0.9367) median parasite densities (4674, 95%CI: 419.0 – 11430 vs 4242, 95%CI: 2436 - 5444 parasites/µL) between the two *pfk13* genotypes (Figure 4). These results suggest that the presence of *pfk13* mutations did not lead to significant differences in parasite density.

**Figure 4.**
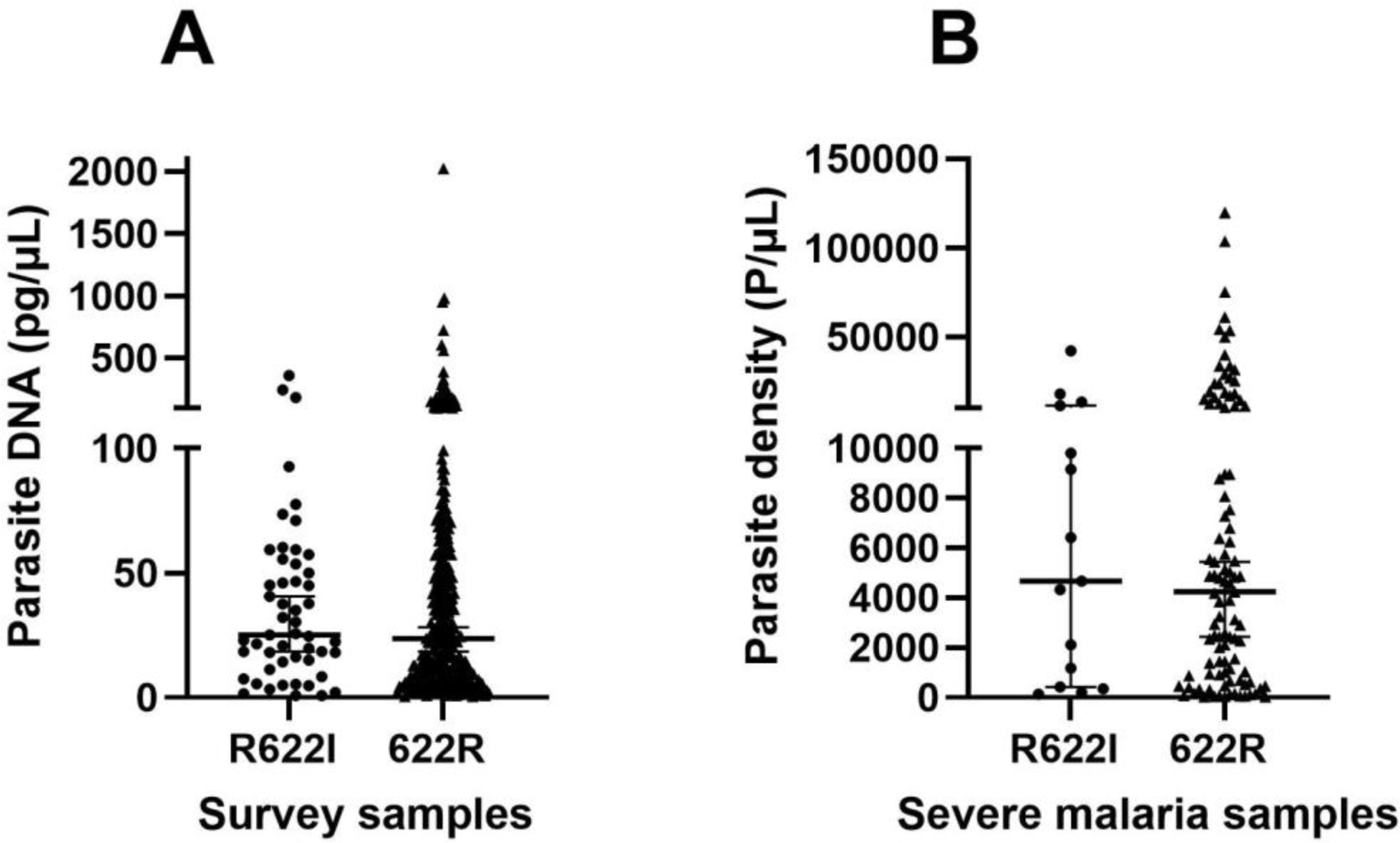
Comparison of parasite density between *pfk13* R622I mutant and wild type 622R parasites. Left panel: Parasite densities in survey participants estimated by qPCR from DBS-derived DNA. Right panel: Parasite densities for participants from the severe malaria study, as estimated from blood films. Horizontal and vertical bars show median density and inter-quartile range.

### Proportion of samples with *pfhrp2/3* deletions

Of the total 587 samples collected between 2018 and 2020 with *pfk13* sequences, 578 had *pfhrp2* and *pfhrp3* status determined. Among these 17 samples (2.9%) displayed a single *pfhrp2* deletion, 255 samples (44.1%) exhibited a single *pfhrp3* deletion, 145 samples (25.1%) with double *pfhrp2/3* deletion, and 161 samples (27.9%) showed no deletion in either *pfhrp2* or *pfhrp3* (Figure 5, Supplemental Table 4). The remaining nine samples could not have their *pfhrp2/3* status determined due to insufficient DNA.

**Figure 5.**
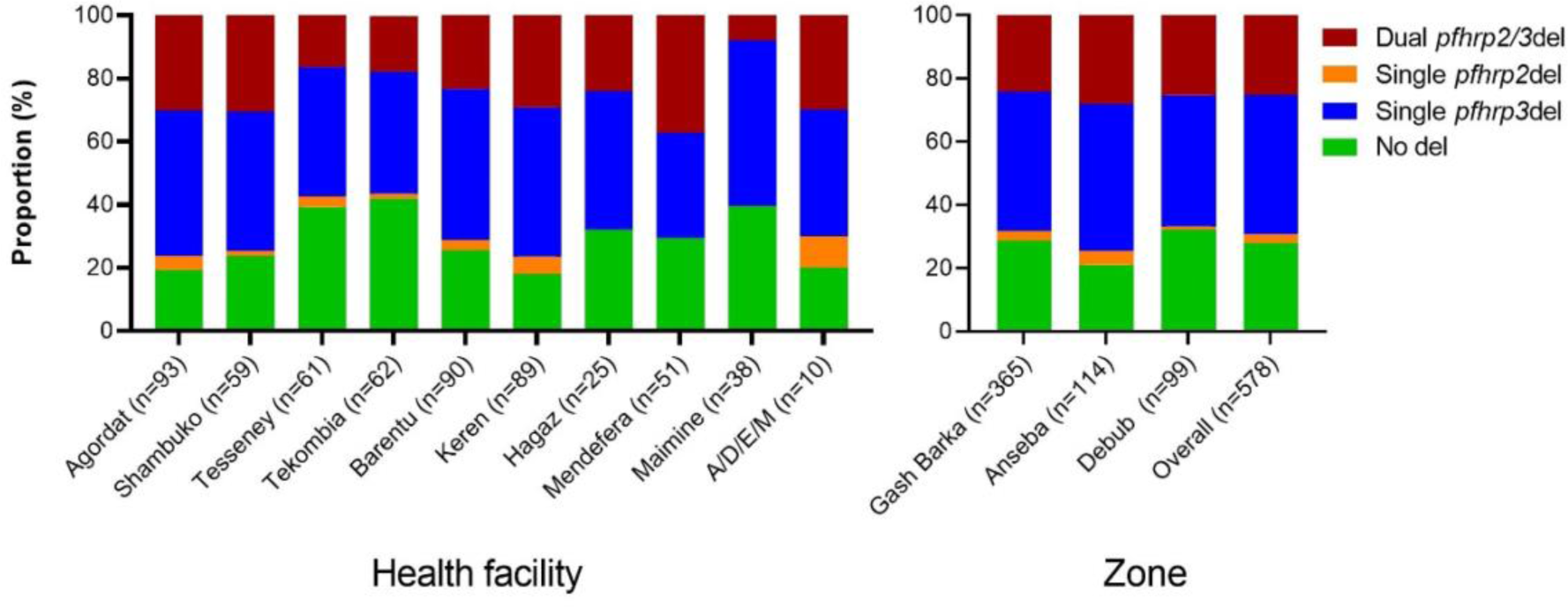
Proportion of samples with and without *pfhrp2* and *pfhrp3* deletions.

### Association of *pfk13* R622I mutation with *pfhrp2/3* deletions

Table 1 provides a summary of *pfk13* mutations observed in samples with varying *pfhrp2/3* status. Samples with single *pfhrp3* deletion, in addition to having four out of five *pfk13* mutations, also showed a significantly higher prevalence of R622I mutation (18.0%, 46/255) compared to samples without *pfhrp2/3* deletions (6.2%, 10/161, OR =3.89, p = 0.0006), or those with dual *pfhrp2/3* deletions (9.0%, 13/145, OR = 2.23, p=0.0183) (Table 1, Supplemental Figure 1A).

**Table 1.** Number and percentage of *pfK13* mutations in samples with varying *pfhrp2* and *pfhrp3* gene status. P-value, confidence interval, CI; and Odds Ratio (OR) are single *pfhrp3* deletion compared to other genotypes.

| *pfhrp2/3* status | No. seq. | E605K |  | R622I |  |  | N657K |  | K658E |  | S679L |  | WT |  |
| --- | --- | --- | --- | --- | --- | --- | --- | --- | --- | --- | --- | --- | --- | --- |
|  |  | n | % | n | % | p-value,[ CI], OR | n | % | n | % | n | % | n | % |
| Single *pfhrp2* del | 17 | 0 | 0.0% | 1 | 5.9% | 0.322, [0.52-150], 3.51 | 0 | 0.0% | 0 | 0.0% | 0 | 0.0% | 16 | 94.1% |
| Single *pfhrp3* del | 255 | 0 | 0.0% | 46 | 18.0% |  | 2 | 0.8% | 1 | 0.4% | 1 | 0.4% | 205 | 80.4% |
| Dual *pfhrp2/3* del | 145 | 0 | 0.0% | 13 | 9.0% | 0.0183,[1.13-4.68], 2.23 | 0 | 0.0% | 0 | 0.0% | 0 | 0.0% | 132 | 91.0% |
| No deletion | 161 | 1 | 0.6% | 10 | 6.2% | 0.0006,[1.59-7.61], 3.89 | 0 | 0.0% | 0 | 0.0% | 0 | 0.0% | 150 | 93.2% |
| Total | 578 | 1 | 0.2% | 70 | 12.1% |  | 2 | 0.3% | 1 | 0.2% | 1 | 0.2% | 503 | 87.0% |

Out of the 70 samples with R622I mutations, one sample (1.4%) was classified as having a single *pfhrp2* deletion, 46 samples (65.7%) showed single *pfhrp3* deletion, 13 samples (18.6%) had dual *pfhrp2/3* deletion and 10 samples (14.3%) had no *pfhrp2/3* deletions (Supplemental Figure 1B). In the remaining 508 wild type 622R samples however, the proportions of single *pfhrp2*, single *pfhrp3*, dual *pfhrp2/3* deletion and no deletions were: 3.1%, 41.1%, 26.0%, and 30.7% respectively (Supplemental Figure 1B). The proportion of single *pfhrp3* deletion was significantly higher (p<0.0001), while the proportion of no deletions (p= 0.0071) was significantly lower in the R622I mutant compared to 622R wild type (Supplemental Figure 1B).

## DISCUSSION

This study reports the prevalence and trends of *pfk13* mutations in the *P. falciparum* population in Eritrea from 2016 to 2020. Additionally, the study explores any potential association between these mutations and *pfhrp2/3* deletions. By analysing these factors, the study aims to gain a better understanding of the underlying factors influencing the prevalence of *pfK13* mutations and *pfhrp2/3* gene deletions.

In 2016, a decade after widespread use of HRP2-based RDTs and AS-AQ in Eritrea, 62% of the patients were infected with parasites lacking both *pfhrp2* and *pfhrp3* genes, rendering them undetectable by HRP2-based RDTs^4^. Notably, no mutation was detected in the *pfk13* gene in these samples. Despite the relatively small sample size, the data suggest that the *pfk13* mutations were either absent or present as a minor variants below the detection limit, indicating very low frequencies at the two study sites during the survey period. This finding aligns with an earlier study conducted in 2013/14, which reported a low prevalence (0.8%, 1/119) of the R622I mutation in samples collected in Agordat and Barentu in the Gash Barka Zone^12^ despite difference in sample collection sites and sequencing technologies. Nevertheless, a more recent study reported a much higher prevalence (8.2%) of R622I mutation among 280 samples collected during the 2016 TES conducted in the Gash Barka Zone. This suggests that the distribution of R622I mutation was highly geographically heterogeneous in Eritrea in 2016.

In 2018-2020, two years after discontinuing the use HRP2-based RDTs, *pfk13* R622I mutations were identified in samples from all health facilities across three zones in Eritrea. The prevalence of this mutation ranged from 2.5% to 28% with a mean prevalence of 11.9%, and more than three-quarters of health facilities reported a prevalence exceeding 10%. In specific locations like Agordat and Barentu, the prevalence of R622I had reached 13.7% (13/95) and 11.0% (10/91), respectively, which is significantly higher than five years earlier^12^. The prevalence and wide distribution of R622I mutation detected in this study combined with that reported recently from TES^10^ demonstrate the R622I mutation has become highly prevalent throughout of Eritrea since 2017. Microsatellite genotyping results revealed that the increase in R622I prevalence was due to a combination of clonal expansion of the mutant parasites and multiple independent emergences from different genetic backgrounds, suggesting a complex evolutionary dynamics similar to that reported in Ethiopia^26^. It is likely that these mutations locally evolved in Eritrea rather than spreading from neighbouring countries such as Sudan and Ethiopia, as genetic analysis using the same haplotype data indicated differences between Eritrean parasite lineages, both with and without *pfhrp2/3* deletions, and those from Sudan and Ethiopia/ Djibouti^27^.

Interestingly, our study found that the R622I mutation was significantly enriched in single *pfhrp3*-deleted (*pfhrp2* positive) parasites and was relatively more prevalent in *pfhrp2/3*-deleted parasites in Eritrea. This observation, which was also reported from TES results^10^ in Eritrea, contrasts with the findings in Ethiopia, where R622I was more prevalent in parasites without *pfhrp2/3* deletions^26^. This difference may be related to the types of RDTs used in these countries at the time of the survey. While both countries had high prevalence of *pfhrp2/3* deletions, Eritrea switched from HRP2-based RDTs to pLDH-based RDTs in 2016, whereas Ethiopia continued to use HRP2-based RDTs. We hypothesise a two-step evolution: an initial emergence of *pfhrp2/3* deletions under HRP2-based RDT selection pressure, followed by the multiple independent emergences of the *pfk13* R622I mutation in gene*-* deleted parasites under ACT selection pressure. In Eritrea, it is likely that *pfhrp2/3* deletion occurred first, given its higher prevalence compared to R622I. However, the driving force for a close association between *pfhrp3* deletion and *pfk13* R622I, and between *pfhrp2/3* deletion and *pfk13* R622I is not clear but may be related to fitness gains in parasites carrying both *pfhrp3* deletion and *pfk13* R622I mutations. While the data suggest that the mutation does not appear to be associated with parasite density in peripheral blood, it could potentially alter other characteristics such as transmissibility.

The influence of *pfhrp2/3* deletions on parasite fitness in infected individuals remains uncertain. Previous studies using genomic data from genetic crosses indicated no fitness cost, which reduces heritability associated with these deletions in the mosquito and liver stages^28^. However, contradictory in vitro results have demonstrated fitness costs^29^, suggesting these may be confined to the blood stages of the life cycle. These findings underscore the need for a deeper understanding of the interplay between drug resistance, *pfhrp2/3* deletions and parasite fitness in the evolution of *P. falciparum*.

The emergence of *P. falciparum* with *pfhrp2/3* deletions and *pfk13* mutations presents a dual-threat scenario, conferring resistance to some diagnostic measures and therapeutic interventions. Indications of potential co-occurrence between *pfhrp2/3* deletions and *pfk13* mutations have been documented, notably in Ethiopia. The findings in this study have shed light on instances of co-occurring *pfk13* R622I and *pfhrp2/3* variants in Eritrea ^26^. At present, a well-established correlation between *pfhrp2/3* deletions and *pfk13* mutations, as well as their collective impact on parasite fitness, remains unclear. *In vitro* experimentations have indicated that the *pfk13* C580Y mutation entails modest fitness costs when accompanied by multicopy *pm2/3*, while it remains fitness-neutral in the presence of single *pm2/3* copies^30^. We hypothesise a comparable scenario of fitness cost interplay between *pfk13* mutations and *pfhrp2/3* deletions. Further studies are required to decipher the interaction between *pfhrp2/3* deletions and *pfk13* mutations.

This study has several notable strengths. Firstly, our study is the first to document the presence of *pfk13* R622I in two administrative zones (Anseba and Debub) and ten sub-zones in addition to zones and sub-zones reported previously (Mihreteab 2023). This observation signifies the expansion of *pfk13* R622I in Eritrean territory. Secondly, it is imperative to highlight that two of the newly identified sub-zones, Tesseney (located in close proximity to the Sudanese border) and Maimine (situated near the Ethiopian border), carry notable regional significance, requiring concerted regional response. Finally, the study’s strength is underscored by the substantial sample size collected in 2019 compared to most recently published ^10^ for the same year. This sample size allows greater power for analysis of association between *pfhrp2/3* deletion and *pfk13* mutation. Overall, our study independently confirms the emergence, progress augmentation overtime and geographical expansion of *pfk13* R622I, along with *pfhrp2/3* deletions correlation within Eritrea context.

This study has certain limitations. Firstly, samples collected before 2016 and after RDT switch (2018-2020) were obtained from different health facilities and zones, limiting direct comparisons in the prevalence of *pfhrp2/3* deletions and *pfk13* mutations between the two time points. Secondly, sequencing of *pfk13* was conducted on a subset of samples collected between 2018 and 2020. However, as all samples from the severe malaria study and a representative subset of samples from the 2019 survey (including 66 – 88% of *pfhrp2/3* deletion and 59% of no deletion samples) were sequenced, significant bias on the outcomes is unlikely. The study by Mihreteab et al. during the same timeframe using samples from TES reported comparable results, thereby confirming the validity of our own findings. Furthermore, this study focused on mutations within the propeller domain of the *pfk13*, excluding possible mutations outside of the propeller domain that may contribute to artemisinin resistance and interaction with gene deletions.

The R622I mutation has primarily been reported in the Horn of Africa region, where very high *pfhrp2/3* deletion prevalence was previously reported^4,6,27,31,32^. Eritrea and Sudan introduced AS-AQ, while Ethiopia and Djibouti use AL as first line therapy for uncomplicated *P. falciparum* malaria. Our study in Eritrea demonstrates the expansion of *pfk13* R622I mutations in the country, with the mutation being enriched in single *pfhrp3*-deleted parasites. While the emergence of *pfk13* R622I is likely associated with increased ACT use and reduced transmission, its interaction with *pfhrp2/3* deletions remains unclear. Although R622I has been established as a confirmed molecular marker of partial artemisinin resistance in Eritrea, additional research is necessary to ascertain its applicability as a molecular marker in neighbouring regions, where *P. falciparum* strains with distinct genetic backgrounds are prevalent. Continued monitoring of the trends in *pfhrp2/3* deletions and *pfK13* R622I mutations in Eritrea and the Horn of Africa with and without HRP2-based RDT uses will provide insight into the relationship between *pfhrp2/3* deletions and *pfK13* R622I mutations, essential for the success of future control and elimination efforts.

## Data Availability

All data produced in the present study are available upon reasonable request to the authors

## Notes

### Acknowledgement

The authors would like to thank patients for their participation of the studies. We would also like to thank Kathleen Krupinski from the WHO GIS Centre for Health for map production.

### Disclaimer

The views expressed in this article are those of the authors and do not necessarily reflect the official policy or position of the Australian Department of Defence, nor the WHO, or the Ministry of Health, Eritrea.

### Author Contributions

SM, KB, JC and QC designed the study. KA, IM, DLF and DS conducted laboratory experiments, acquired data and performed initial data analysis. CS contributed materials and data analysis. SM organised and managed field surveys, sample collection and epidemiological analysis. KB and QC analysed data and drafted the manuscript. All authors participated in manuscript review and revision, and approved the final manuscript.

### Author contact details

Selam Mihreteab

Karen Anderson

Irene Molina - de la Fuente

Colin J Sutherland

David Smith

Jane Cunningham

Khalid B Beshir

Qin Cheng

