## Supplementary materials for "The spread of a validated molecular marker of artemisinin partial resistance *pfkelch13* R622I and association with *pfhrp2/3* deletions in Eritrea"

[1]National Malaria Control Program, Ministry of Health, Asmara, Eritrea; [2]Drug Resistance and Diagnostics, Australian Defence Force Malaria and Infectious Disease Institute, Brisbane, Australia; [3]QIMR-Berghofer Medical Research Institute, Brisbane, Australia; [4] Carlos III Institute of Health, Madrid, Spain; [5]University of Alcala, Madrid, Spain; [6]Faculty of Infectious Diseases, London School of Hygiene & Tropical Medicine, UK; [7]Global Malaria Programme, World Health Organization, Geneva, Switzerland.

*Corresponding authors

Dr Qin Cheng, Australian Defence Force Malaria and Infectious Disease Institute, Weary Dunlop Drive, Gallipoli Barracks, Enoggera, QLD 4051, Australia..

Dr Khalid B Beshir, Faculty of Infectious Diseases, London School of Hygiene & Tropical Medicine, UK..

**Supplemental Table 1**. Number of samples sequenced for *pfk13*, mutations identified and mutation frequency in different health facilities.

| Zone | Health Centre | No. sequenced | E605K | | R622I | | N657K | | K658E | | S679L | | WT | |
|---|---|---|---|---|---|---|---|---|---|---|---|---|---|---|
| | | | n | % | n | % | n | % | n | % | n | % | n | % |
| Gash Barka | Agordat | 95 | 0 | 0.0% | 13 | 13.7% | 1 | 1.1% | 0 | 0.0% | 0 | 0.0% | 81 | 85.3% |
| | Shambuko | 61 | 1 | 1.6% | 7 | 11.5% | 0 | 0.0% | 1 | 1.6% | 0 | 0.0% | 52 | 85.2% |
| | Tesseney | 61 | 0 | 0.0% | 7 | 11.5% | 0 | 0.0% | 0 | 0.0% | 0 | 0.0% | 54 | 88.5% |
| | Tokombia | 62 | 0 | 0.0% | 7 | 11.3% | 0 | 0.0% | 0 | 0.0% | 0 | 0.0% | 55 | 88.7% |
| | Barentu | 91 | 0 | 0.0% | 10 | 11.0% | 1 | 1.1% | 0 | 0.0% | 0 | 0.0% | 80 | 87.9% |
| | *Subtotal* | *370* | *1* | *0.3%* | *44* | *11.9%* | *2* | *0.5%* | *1* | *0.3%* | *0* | *0.0%* | *322* | *87.0%* |
| Anseba | Keren | 91 | 0 | 0.0% | 9 | 9.9% | 0 | 0.0% | 0 | 0.0% | 0 | 0.0% | 82 | 90.1% |
| | Hagaz | 25 | 0 | 0.0% | 7 | 28.0% | 0 | 0.0% | 0 | 0.0% | 0 | 0.0% | 18 | 72.0% |
| | *Subtotal* | *116* | *0* | *0.0%* | *16* | *13.8%* | *0* | *0.0%* | *0* | *0.0%* | *0* | *0.0%* | *100* | *86.2%* |
| Debub | Mendefera | 51 | 0 | 0.0% | 3 | 5.9% | 0 | 0.0% | 0 | 0.0% | 0 | 0.0% | 48 | 94.1% |
| | Maymine | 40 | 0 | 0.0% | 5 | 12.5% | 0 | 0.0% | 0 | 0.0% | 1 | 2.5% | 34 | 85.0% |
| | A/D/E/M | 10 | 0 | 0.0% | 2 | 20.0% | 0 | 0.0% | 0 | 0.0% | 0 | 0.0% | 8 | 80.0% |
| | *Subtotal* | *101* | *0* | *0.0%* | *10* | *9.9%* | *0* | *0.0%* | *0* | *0.0%* | *1* | *1.0%* | *90* | *89.1%* |
| **Total** | | **587** | **1** | **0.2%** | **70** | **11.9%** | **2** | **0.3%** | **1** | **0.2%** | **1** | **0.2%** | **512** | **87.2%** |

**Supplemental Table 2**. Haplotypes observed among 26 samples carrying *pfk13* R622I mutation.

| Zone | Location | SAMPLE ID | TA1 | 313 | 2490 | 383 | POLY A | PK2 | TA109 | Haplo type |
|---|---|---|---|---|---|---|---|---|---|---|
| Gash Barka | Agordat | EA04 | 165 | 229 | 83 | 137 | 152 | 162 | 160 | 1 |
| | Agordat | EA02 | 165 | 233 | 83 | 137 | 152 | 162 | 160 | 2 |
| | Agordat | EA28 | 165 | 233 | 83 | 137 | 152 | 162 | 160 | 2 |
| | Agordat | EA66 | 165 | 233 | 83 | 137 | 158 | 162 | 151 | 3 |
| | Barentu | EB1988 | 159 | 253 | 83 | 125 | 152 | 162 | 172 | 4 |
| | Barentu | EB2254 | 165 | 219 | 83 | 125 | 158 | 162 | 160 | 5 |
| | Barentu | EB1793 | 165 | 219 | 83 | 127 | 158 | 171 | 160 | 6 |
| | Barentu | EB1986 | 165 | 237 | 83 | 127 | 167 | 171 | 175 | 7 |
| | Shambuko | ES65 | 165 | 233 | 83 | 137 | 152 | 162 | 151 | 8 |
| | Shambuko | ES26 | 165 | 233 | 83 | 137 | 152 | 171 | 151 | 9 |
| | Shambuko | ES61 | 165 | 235 | 83 | 137 | 158 | 162 | 175 | 10 |
| | Tesseney | ETHOS03 | 165 | 229 | 83 | 137 | 158 | 162 | 151 | 11 |
| | Tesseney | ETHOS30 | 165 | 247 | 83 | 136 | 152 | 162 | 154 | 12 |
| | Tokombia | ETHC59 | 159 | 229 | 83 | 137 | 152 | 162 | 151 | 12 |
| | Tokombia | ETHC60 | 165 | 233 | 83 | 137 | 173 | 162 | 175 | 14 |
| | Tokombia | ETHC42 | 165 | 235 | 83 | 137 | 158 | 162 | 165 | 15 |
| Anseba | Hagaz | EH07 | 165 | 229 | 83 | 137 | 149 | 162 | 160 | 16 |
| | Hagaz | EH23 | 165 | 233 | 83 | 127 | 167 | 171 | 160 | 17 |
| | Hagaz | EH04 | 165 | 233 | 83 | 137 | 170 | 162 | 163 | 18 |
| | Hagaz | EH20 | 165 | 233 | 83 | 137 | 173 | 162 | 175 | 14 |
| | Hagaz | EH29 | 165 | 233 | 83 | 137 | 173 | 162 | 175 | 14 |
| | Hagaz | EH17 | 165 | 249 | 83 | 137 | 152 | 162 | 163 | 19 |
| | Keren | EK67 | 165 | 233 | 83 | 137 | 152 | 162 | 163 | 20 |
| Debub | Maimine | EMA02 | 159 | 233 | 83 | 127 | 176 | 186 | 172 | 21 |
| | Maimine | EMA33 | 165 | 229 | 83 | 137 | 155 | 186 | 175 | 22 |
| | Mendefera | EME31 | 165 | 229 | 83 | 137 | 158 | 162 | 160 | 23 |

**Supplemental Table 3**. Number of unique alleles and genetic diversity detected per microsatellite locus and population.

| Microsatellite marker | Number of alleles | | Gene diversity $H_E$ per locus | |
|---|---|---|---|---|
| | Mutant | Wild type | Mutant | Wild type |
| TA1 | 2 | 8 | 0.212 | 0.647 |
| 313 | 8 | 8 | 0.745 | 0.542 |
| 2490 | 1 | 5 | 0.000 | 0.167 |
| 383 | 3 | 9 | 0.394 | 0.455 |
| PolyA | 8 | 13 | 0.785 | 0.828 |
| PK2 | 3 | 9 | 0.394 | 0.674 |
| TA109 | 7 | 10 | 0.812 | 0.728 |
| Total | 23 | 87 | Mean (95% CI)<br>0.4774 (0.1873 - 0.7676) | Mean (95% CI)<br>0.5773 (0.3760 – 0.7786) |
| | | | P = 0.0500 | |

**Supplemental Table 4.** Number and proportion of samples with different *pfhrp2/3* status.

| Zone | Health Centre | No. Tested | Single *pfhrp2* del | | Single *pfhrp3* del | | Dual *pfhrp2/3* del | | No del | |
|---|---|---|---|---|---|---|---|---|---|---|
| | | | n | % | n | % | n | % | n | % |
| Gash Barka | Agordat | 93 | 4 | 4.3% | 43 | 46.2% | 28 | 30.1% | 18 | 19.4% |
| | Shambuko | 59 | 1 | 1.7% | 26 | 44.1% | 18 | 30.5% | 14 | 23.7% |
| | Tesseney | 61 | 2 | 3.3% | 25 | 41.0% | 10 | 16.4% | 24 | 39.3% |
| | Tokombia | 62 | 1 | 1.6% | 24 | 38.7% | 11 | 17.7% | 26 | 41.9% |
| | Barentu | 90 | 3 | 3.3% | 43 | 47.8% | 21 | 23.3% | 23 | 25.6% |
| | *Subtotal* | *365* | *11* | *3.0%* | *161* | *44.1%* | *88* | *24.1%* | *105* | *28.8%* |
| Anseba | Keren | 89 | 5 | 5.6% | 42 | 47.2% | 26 | 29.2% | 16 | 18.0% |
| | Hagaz | 25 | 0 | 0.0% | 11 | 44.0% | 6 | 24.0% | 8 | 32.0% |
| | *Subtotal* | *114* | *5* | *4.4%* | *53* | *46.5%* | *32* | *28.1%* | *24* | *21.1%* |
| Debub | Mendefera | 51 | 0 | 0.0% | 17 | 33.3% | 19 | 37.3% | 15 | 29.4% |
| | Maimine | 38 | 0 | 0.0% | 20 | 52.6% | 3 | 7.9% | 15 | 39.5% |
| | A/D/E/M | 10 | 1 | 10.0% | 4 | 40.0% | 3 | 30.0% | 2 | 20.0% |
| | *Subtotal* | *99* | *1* | *1.0%* | *41* | *41.4%* | *25* | *25.3%* | *32* | *32.3%* |
| **Total** | | **578** | **17** | **2.9%** | **255** | **44.1%** | **145** | **25.1%** | **161** | **27.9%** |

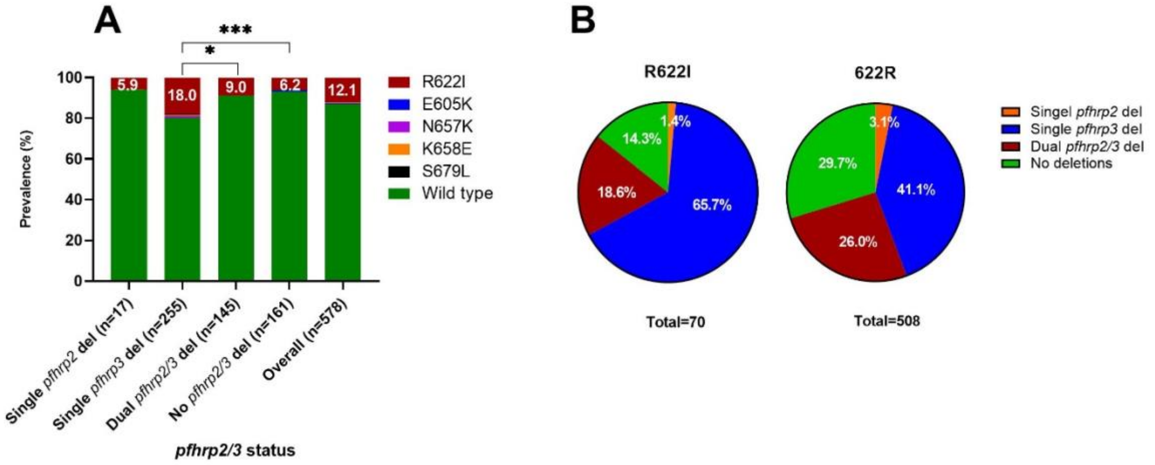


**Supplementary Figure 1**. Prevalence K13 mutations in samples with different *pfhrp2/3* status (A) and proportion of *pfhrp2/3*-deleted parasites among K13 R622I mutant and 622R wild type (B).
